# Multiparametric MRI detects multi-organ impairment in patients with chronic myeloid neoplasms with normal serum biomarkers

**DOI:** 10.1101/2023.11.23.23298558

**Authors:** Sophie Reed, Charlie Diamond, Samiya Mahmood, Soubera Rymell, Michael Smith, Michele Pansini, Bethan Psaila, Adam J. Mead, Helena Thomaides-Brears, Onima Chowdhury

**Affiliations:** Oxford University Hospitals’ NHS Foundation Trust, Oxford, UK; Molecular Haematology Unit, Weatherall Institute of Molecular Medicine, NIHR, Biomedical Research Centre, University of Oxford, Oxford, UK; Perspectum Ltd, Gemini One, 5520 John Smith Drive, Oxford, UK

## Abstract

Assessment of organ impairment in patients with chronic myeloid neoplasms is pivotal in selecting treatments and for accurate prognostication of patient outcomes. In order to determine the multi-organ health of patients with chronic myeloid neoplasms, we conducted a prospective, observational study utilising a novel MRI technology which quantitatively assesses the health of multiple organs in one scan. Organ impairment was significantly higher in the patient cohort compared to healthy controls, most notably with increased rates of kidney fibroinflammation 28% vs 0% (p-value = 0.002). MRI-defined kidney impairment was prevalent in patients with normal serum biomarkers of kidney disease, demonstrating the added value of MRI as a tool to identify occult organ impairment. This has wider implications for enhancing the assessment of organ health in patients with a variety of blood cancers at diagnosis and throughout treatment, guiding more personalised strategies and improving patient outcomes.

## Introduction

Organ impairment and organomegaly are key features of chronic phase myeloid neoplasms and impact significantly on clinical decision-making. In the *BCR ABL* negative myeloproliferative neoplasms (MPNs), extramedullary haematopoiesis, inflammation and fibrosis all play a role in disease biology and patient outcomes (1). Kidney impairment has specifically been reported to correlate with poor outcome, particularly in patients with essential thrombocythemia (ET) (2). In patients with chronic myeloid leukaemia (CML), pre-existing cardiac, liver or renal dysfunction strongly influence the choice of tyrosine kinase inhibitor (TKI) (3,4). Additionally, patient comorbidities have been shown to be the most powerful predictor of overall survival in patients with CML, regardless of TKI response, demonstrating the importance of carefully assessing organ health at diagnosis(5). Autoimmune and inflammatory conditions have been reported in up to 30% of patients with myelodysplastic syndromes (MDS) (6) where multi-organ consequences may not be recognised as readily as in the MPNs. Sensitive and reproducible assessment of organ impairment is central to optimal stratification, prognostication and treatment selection in these patient groups. Currently, assessment of organ impairment is broadly reliant on blood tests, ultrasound/CT based imaging or invasive biopsies. These have a low specificity and sensitivity, and can present a clinical risk to the patients in the case of biopsies. An integrated non-invasive approach for early identification of organ impairment, which could be used serially, may present a more effective pathway for patients with chronic myeloid neoplasms, and be particularly valuable prior to high intensity treatment modalities such as stem cell transplantation. CoverScan is a UKCA (UK conformity assessed) marked quantitative magnetic resonance imaging (MRI) analysis which can determine the health of multiple organs in a single scan, using existing conventional MR systems. CoverScan is superior to standard MR techniques as in addition to analysing organ size, it enables *in vivo* quantitative tissue characterisation of features such as fibroinflammation and fat content, in multiple organs in one forty minute scan. It has been utilised to detect organ damage in multi-organ diseases such as Long Covid and Diabetes (7–9). We sought to determine multi-organ health in patients with chronic myeloid neoplasms, including CML, MDS and MPNs, utilising Coverscan and blood biomarkers. Integrating quantitative imaging assessment into routine testing, could help to refine this approach and improve decision making. To our knowledge, this is the first study of the utility of multi-organ imaging by MRI in patients with any blood cancers.

## Methods

We conducted a single centre, prospective, longitudinal, observational study of patients with chronic myeloid neoplasms, who had recovered from COVID-19 (MyeloidScan REC 292188). Participants were eligible for the study if they had a chronic myeloid neoplasm and were able to undergo MRI scanning. Using a combination of CoverScan MRI and blood biomarkers of inflammation, organ function and immune response, we set out to determine the degree of multi-organ impairment at two timepoints 6-months apart. Matched controls (age and sex) were retrospectively selected from amongst 92 healthy controls (Supplementary Methods). Study IDs were not known to anyone outside the research group.

### MRI Definition of organ impairment

CoverScan provides organ-specific measures of size, fat deposition, and fibroinflammation. T1 weighted MRI quantifies free water content in biological tissues and can be diagnostic of disease histology such as inflammation or fibrosis. MRI-derived measurements from the heart, lungs, kidneys, liver, pancreas and spleen were compared with established reference ranges to determine impairment for each organ. Organ impairment was defined for each metric according to established reference ranges from healthy controls (Table S1) and was grouped by evidence of: borderline or low ejection fraction and evidence of myocarditis in the heart; reduced pulmonary dynamic measurements in the lungs; elevated cortical T1 in the kidneys; steatosis or inflammation in the liver and pancreas; and splenomegaly. Multi-organ impairment at baseline and follow-up was defined as ≥2 MRI measurements from different organs outside reference ranges. Further details including statistical analysis are available in Supplementary Methods.

## Results

Twenty six patients with chronic myeloid neoplasms were enrolled between May 2021 and July 2022 (MPN, *n* = 17 [polycythaemia (PV), *n* = 5; ET, *n* = 9; myelofibrosis (MF), *n* = 3]; CML, *n* = 7; MDS *n* = 2). At the baseline scan, the mean (SD) age of the patients was 55 years (13), with an equal split between male and females (Table S2). The mean (SD) age of the healthy control cohort (*n* = 35) was 52 years (7) and the percentage of females was 66%. 58% (15/26 patients) were on active therapies, including hydroxycarbamide (*n* = 4), pegylated interferon (*n* = 2), and TKIs (imatinib *n* = 2; dasatinib *n* = 2; nilotinib *n* = 2; bosutinib *n* = 1). One MDS patient had developed secondary MDS, post PV and was receiving azacitidine, the other had primary low risk MDS treated with darbepoietin. 42% of all patients (11/26) were not receiving any treatment and were being actively monitored. Comorbidities and blood test results from study enrolment and follow-up are reported (Table S3). All patients with CML had a standard *BCR ABL* translocation at diagnosis and were in major molecular remission (MMR) at the time of study enrolment. All patients with an MPN had a *JAK2, CALR* or *MPL* driver mutation.

Organ impairment was substantially higher in patients with chronic myeloid neoplasms when compared to the healthy controls: kidney fibroinflammation (28% vs 0% p-value = 0.002), liver fibroinflammation (20% vs 5.7% p-value = 0.12), pancreatic steatosis or fibroinflammation (38% vs 17% p-value = 0.061) (Figure 1). At least 1 organ was impaired in 73% vs 29% (p-value <0.001) of patients and healthy controls, with multi-organ impairment (≥ 2 organ impaired) being observed in 42% vs 5.7% (p-value <0.001) respectively. The most striking and unexpected impairment was in the kidney. Of the 7 patients with an abnormal baseline kidney MRI result (ET, *n* = 3; MF, *n* = 2; CML, *n* = 1; MDS, *n* = 1), only 1 patient (diagnosis MF) had a corresponding elevated serum creatinine and low estimated glomerular filtration rate (eGFR) of 56.9 ml/min/1.73m^2^ (Figure 2, Table S4). Focussing on the patients with kidney impairment in more detail, the most frequent diagnosis was ET (3/7). These 3 patients were all women aged <55 years old, 2 of which had no additional comorbidities and were treated with aspirin alone, whilst the third was treated with hydroxycarbamide due a previous coronary artery thrombus but had no other comorbidities. The two patients with MF and kidney fibroinflammation, had distinct clinical phenotypes. One patient had low risk MF and was on active monitoring but did have comorbidities including a significantly raised BMI and hypertension. The second patient developed secondary MF, approximately 30 years after being diagnosed with PV treated with hydroxycarbamide and was imminently due start Ruxolitinib but had no other significant comorbidities. This was the only patient in the cohort with an elevated serum creatinine and low eGFR. The only patient with CML and baseline kidney impairment on MRI had no significant comorbidities at the time but subsequently developed breast cancer within 6 months of study enrolment. Twenty one patients attended for a follow-up scan, which identified persisting kidney impairment in all 7 patients with kidney impairment at baseline. At follow-up, there were also 4 new cases with kidney fibroinflammation (ET *n* = 3 and CML *n* = 1), all with normal serum creatine and eGFR at both baseline and follow-up. Reviewing any changes in clinical and/or laboratory parameters between the two scans identified one patient who had recently suffered a cardiac event resulting in occlusion and subsequent stenting of the coronary arteries, approximately 3 months prior to the follow-up MRI scan. However the cardiac component of the follow-up CoverScan was within normal limits. There were no notable changes in underlying myeloid neoplasm, therapeutics, or new comorbidities in the remaining 3 participants, which might explain the new kidney fibroinflammation.

**Figure 1:**
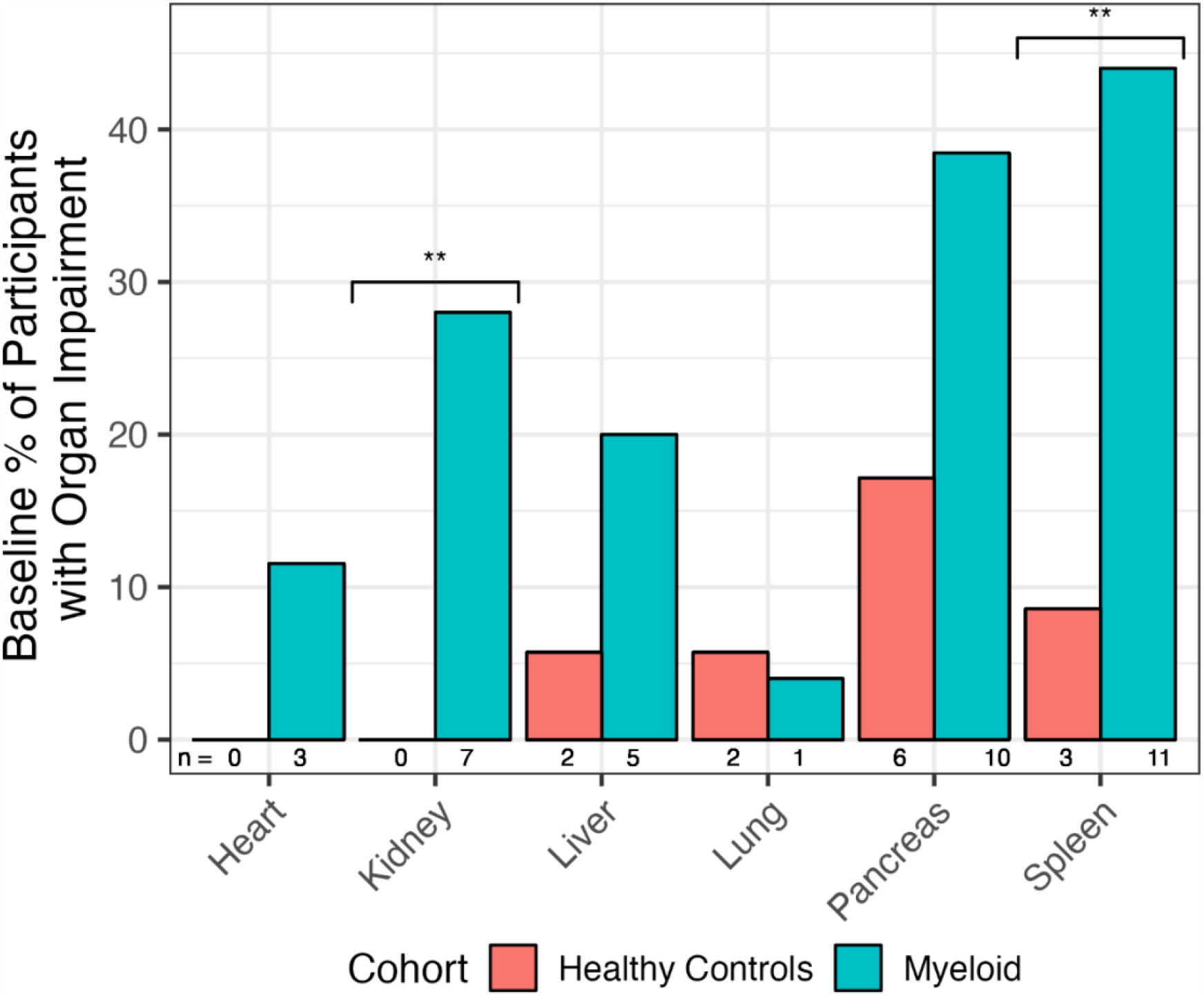
Frequency of participants with MRI defined organ impairment at baseline scan. Bar chart represents total MyeloidScan cohort with chronic myeloid neoplasms (green n = 26) versus healthy controls (red n = 35). ** <p<0.01.

**Figure 2:**
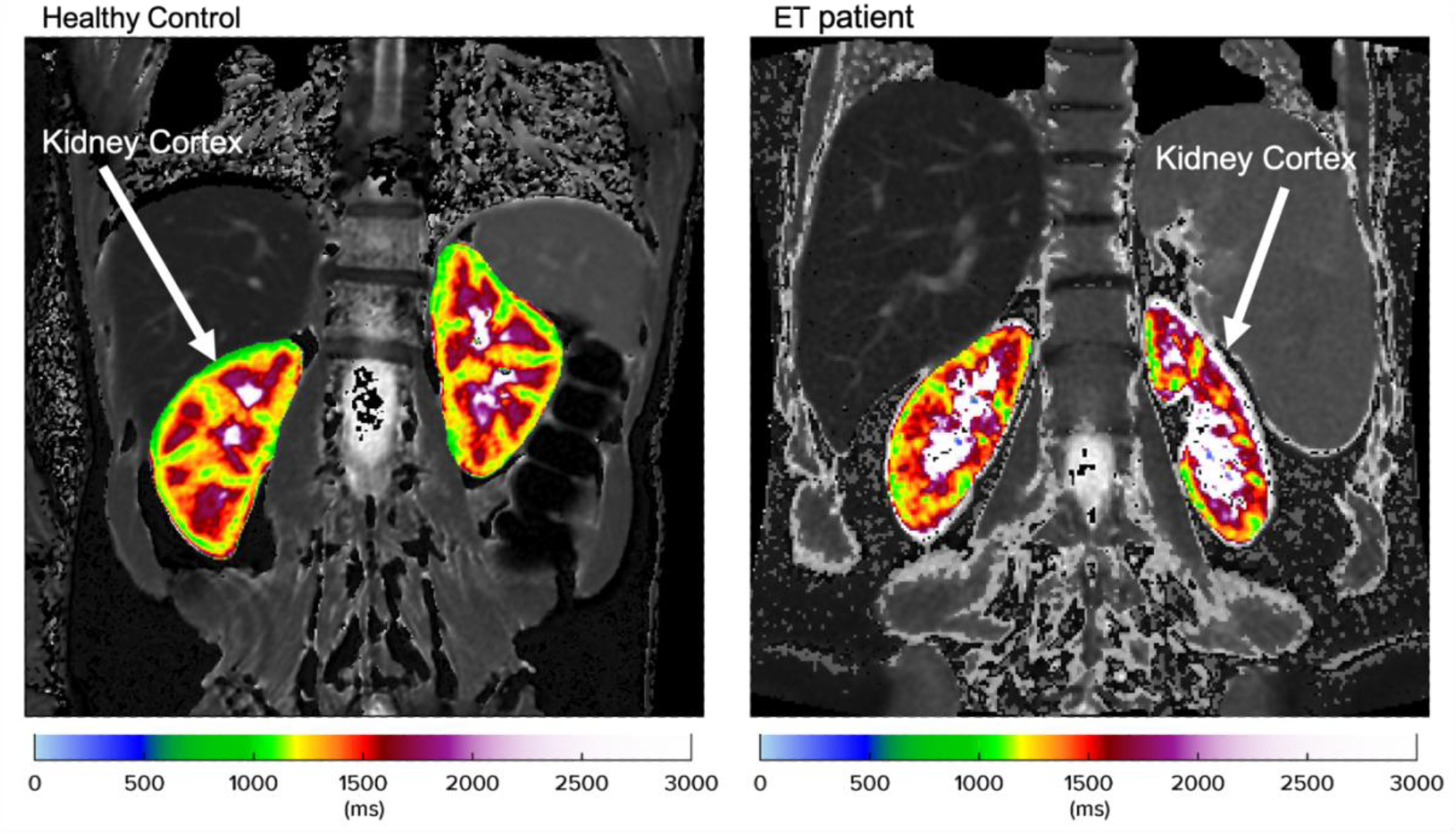
Example MRI Cortical T1 mapping image of normal kidneys from a healthy control (left, green signal in cortex represents T1 within normal range as per heatmap scale.) and abnormal kidneys with an elevated cortical T1 in an example MyeloidScan participant (right, with elevated cortical T1 as per heatmap scale). Healthy: left kidney cortex = 1143ms (ref: <1185ms), right kidney cortex = 1133ms (ref: <1173ms). ET patient: left kidney cortex = 1243ms (ref: <1185ms); right kidney cortex = 1183ms (ref: <1173ms).

## Discussion

CoverScan MRI has revealed a high prevalence of organ impairment in patients with chronic myeloid neoplasms compared to healthy controls. This multi-organ MRI platform offers the capacity to accurately evaluate organ health dynamically. Early detection of such asymptomatic impairment is likely to be informative with regards to long term treatment selection and potentially patient outcomes. Due to the limitations of our study design and numbers, it is not possible to clearly distinguish between myeloid neoplasm-associated end organ damage, treatment related effects or co-morbidities. Regardless of the aetiology it is clear that organ impairment, as determined by validated comorbidity indexes, has a strong impact on overall survival (5). Amongst the patient subgroups, it was striking that renal impairment was evident on MRI in patients with normal serum biomarkers such as creatinine and eGFR. Previous studies in patients without blood cancers demonstrate that cortical T1 is highly predictive of renal outcomes: cortical T1 negatively correlates with kidney function measured by eGFR (10), positively correlates with interstitial fibrosis (11) in chronic kidney disease (CKD), is elevated in CKD and after acute kidney injury (12), and accurately diagnoses and stages diabetic nephropathy (13). This suggests the possibility of using quantitative MRI as an early detection tool for fibroinflammatory disease in patients with myeloid neoplasms. The observed discordance between normal serum biomarkers such as creatinine and eGFR, and the presence of renal impairment as detected by MRI underscores the potential utility of this imaging technique in revealing subclinical pathology that is often missed by conventional testing. The clinical implications at this stage are uncertain, but as renal function is a prognostic factor in MPN, examination of larger patient groups is warranted (14,15). Further studies incorporating multiorgan imaging biomarkers into diagnostic and stratification pathways are required. This could be readily implemented into clinical trial pathways, particularly as patients with MPNs in particular, often already have serial standard MRI scans to assess spleen size. The application of a multi-organ MRI platform like CoverScan could help in the early detection and longitudinal monitoring of organ impairments in patients with chronic myeloid neoplasms, potentially guiding more personalised treatment strategies and improving patient outcomes. With the advent of new therapeutic options on the horizon, identifying organ damage in patients with myeloid neoplasms will likely impact on clinical trial recruitment as well as patient outcomes.

## Data Availability

All data produced in the present study are available upon reasonable request to the authors.

## Author Contributions

OC designed the study, performed the research and wrote the manuscript. SR performed the research, analysed data and wrote the manuscript. CD and HTB analysed data and wrote the manuscript. SM, SR, MS and MP performed research. BP and AJM critically revised the manuscript.

## Acknowledgments

The study was supported by the MRC and CRUK. The authors would like to thank all the patients and patient support groups (MPN Voice, MDS UK and CML Support) for their invaluable support.

