## Supplementary Information for "Multiparametric MRI detects multi-organ impairment in patients with chronic myeloid neoplasms with normal serum biomarkers"


| Page | Content |
|---|---|
| 2 | Supplementary Methods |
| 4 | Table S1: Reference range thresholds for MRI metrics of organ impairment |
| 5 | Table S2: Baseline characteristics of myeloid cancer patients vs healthy controls |
| 7 | Table S3: Clinical characteristics for myeloid cancer patients |
| 10 | Table S4: Characteristics of myeloid cancer patients with kidney fibroinflammation |
| 13 | Supplementary references |

### Supplementary methods

##### Inclusion and exclusion criteria

Inclusion criteria were adults with a myeloid cancer diagnosis, who were able to consent and attend the MRI facility for up to three visit. Exclusion criteria were being cured of blood cancer, contraindications to MRI and any active COVID-19 infection symptoms.

##### Study recruitment

Quantitative multi-organ MRI (CoverScan, Perspectum, Oxford) was used to assess organ impairment as previously reported [1]. Participants underwent a 40-minute MRI scan of lungs, heart, kidney, liver, pancreas and spleen on 1.5T Siemens scanners at the Oxford Community Diagnostics centre on a MAGNETOM Aera 1.5T or a SIGNA Voyager 1.5T. MR metrics were standardised to deliver a single report interpretable by clinicians. Each report included 49 organ-specific metrics with reference ranges to determine impairment. Matched controls (age and sex) were retrospectively selected from amongst 92 healthy controls (no prior COVID-19 diagnosis, no hospital discharge ≥4 months prior to enrolment and contraindications to MRI) who underwent CoverScan as part of an earlier prospective study of largely non-hospitalised long COVID patients (NCT04369807) [1].

##### Imaging Acquisitions

- Cardiac MR imaging involved complete coverage of the heart with a short-axis stack (from the apex to the valve plane) of cine images acquired using cardiac gating, each acquired within a short breath-hold. This acquisition mirrors the one used at the UK Biobank and is a standardized approach [2]. Three short-axis and two long axis cine (HLA and VLA) cardiac T1 maps were also acquired using the MOLLI-T1 approach at the basal, mid, and apical levels of the left ventricle. Three short-axis and two long axis cine (HLA and VLA) at the basal, mid, and apical levels. For cardiac T2 maps, acquired using the fast low angle shot (FLASH) in 3T, at 1.5T scanner, TrueFISP was used (a Siemens version of balanced SSFP).
- Liver and pancreas imaging used the LiverMultiScan acquisition protocol (Perspectum, Oxford, UK), which involves 3 single 2D axial slice breath-held acquisitions that separately are sensitive to the fat content (proton density fat fraction [PDFF]), to T2* (which can yield liver iron content) and a MOLLI-T1 measurement (providing a measurement of tissue water) [3].
- Lungs: Two single slice dynamic cine MR acquisitions were acquired in the coronal plane with a 307ms temporal resolution: one 40 s acquisition with the patient instructed to breathe normally and a second 30 s acquisition with the patient instructed to breathe deeply.
- Kidney: a single coronal view was used to image both kidneys. Imaging contrasts were MOLLI-T1, and a spoiled gradient recalled acquisition (spGR).
- Spleen: Volumetric 3D spGR MRI images for organ length. LiverMultiScan acquisition protocol above was used for spleen T1.

##### Image Analysis

- Cardiac: Experienced cardiac MRI analysts used CVI42v5.11 (Cardiovascular Imaging Inc, Canada) to trace manually the myocardium in the end-diastolic and end-systolic phases in each of the short-axis views, following the standard UK Biobank evaluation approach as previously described [4]. We reported ventricular function; end systolic and diastolic volume; stroke volume and ejection fraction in both ventricles; left ventricular muscle mass and ventricular max wall thickness and global longitudinal and circumferential 3D strain metrics. Cardiac T1 and T2 measurements were determined as the mean of each of the 16 cardiac segments (of the AHA 17 segment model excluding the apex) [5]. When artefacts were present those measurements were not included in the results.
- Liver Images were analysed by data analysts experienced at using the LiverMultiScan (Perspectum, Oxford, UK) software. This yielded global metrics in each liver of PDFF (proton density fat fraction),

T2*, and cT1 (cT1 is a measurement of T1 that has been corrected for the confounding effects of iron and standardised to 3 Tesla; it is elevated with disease).

- Pancreas images were analysed in an equivalent manner to the above except the software used was not FDA-cleared and iron correction was not performed [6]. The T1 was standardized to 3 Tesla.
- Lung: Patient respiration was assessed by imaging a single 2D coronal slice of the lungs using a 30 second dynamic cine MRI acquisition (with a time resolution of 307ms), during which the patient was instructed to breathe deeply. In-house developed automated segmentation methods were used to segment the lungs and measure their areas in each time frame. Segmentations were reviewed and corrected by trained analysts. From these area vs time measurements, the lung area at maximum inspiration and expiration was measured, as well as a fractional area change (max area - min area)/max area.
- Kidney: assessed using in-house tools to fit parametric maps and to allow trained analysts to make measurements. The kidney cortex was manually segmented using the MOLLI-T1 map to guide the boundary. Multiple regions-of-interests were manually placed within the cortex to extract a median value of cortical T1 in each kidney. Volumetric delineations of the kidneys were derived from SPGR MRI images. Automated delineations were produced using a 3D convolutional neural network, trained on expert annotations. Delineations were manually checked, and corrected, if necessary, for each subject.
- Spleen: Volumetric delineations were derived from SPGR MRI images. Automated delineations were produced using a 3D convolutional neural network, trained on expert annotations. Delineations were manually checked, and corrected, if necessary, for each subject. Spleen T1 images were analysed in an equivalent manner to liver cT1 and pancreas srT1 except the software used was not FDA-cleared and iron correction was not performed.

**Reference ranges**

All MRI metric reference ranges, but organ volumes, were calculated retrospectively from n=92 healthy controls scanned at 1.5T and 3T based on the 2.5% (lower threshold) and 97.5% percentiles (upper threshold), using bootstrapping (100,000 permutations) [1]. Spleen volumes were calculated from a combined cohort of 92 healthy controls and 1744 BMI-matched participants from the UK Biobank [7], representing all sex and height subgroups, as these are known confounders of organ size [8].

**Organ Impairment**

Organ impairment was calculated for each organ based on evidence of any of the measurements appearing out of reference range (Supplementary Table 1, essentially as in [1]). Thus, depending on the metric the upper or lower threshold was considered. For cardiac T1 or cardiac T2, elevation in at least 3 AHA segments defined elevated T1. Single organ impairment was based on at least 1 organ impaired and multi-organ impairment was based on 2 organ impairments.

**Statistical analysis**

We presented the descriptive statistics for continuous and categorical variables with the mean (standard deviation, SD) and frequency (percentage), respectively. For groupwise comparisons, we used Wilcoxon rank sum tests for continuous variables and Fishers exact test for categorical variables. Statistical significance was defined by a p-value threshold of $<0.05$ (2-sided). All data cleaning and analyses were conducted in R software version 4.2.1.

Table S1: Reference range thresholds for MRI metrics of organ impairment

<table>
<tr><th>Metric</th><th>Reference range</th><th>Definition of impairment</th></tr>
<tr><td>Liver</td><td></td><td></td></tr>
<tr><td>Liver cT1 (ms)</td><td>< 800ms</td><td rowspan="2">Liver cT1 abnormal AND/OR Liver PDFF abnormal</td></tr>
<tr><td>Liver PDFF (%)</td><td>< 5%</td></tr>
<tr><td>Pancreas</td><td></td><td></td></tr>
<tr><td>Pancreas srT1 (ms)</td><td>< 836ms</td><td rowspan="2">Pancreas srT1 abnormal AND/OR Pancreas PDFF abnormal</td></tr>
<tr><td>Pancreas PDFF (%)</td><td>< 4%</td></tr>
<tr><td>Heart</td><td></td><td></td></tr>
<tr><td>Cardiac T1 segment (ms)</td><td>< 1038ms</td><td rowspan="4">(≥3 cardiac T1 segments abnormal AND ≥3 cardiac segments T2 abnormal)<br>OR<br>(≥3 cardiac T1 segments abnormal AND [Left ventricular ejection fraction abnormal OR Right ventricular ejection fraction abnormal])<br>OR<br>(≥3 cardiac T2 segments abnormal AND [Left ventricular ejection fraction abnormal OR Right ventricular ejection fraction abnormal])</td></tr>
<tr><td>Cardiac T2 segment (ms)</td><td>< 58ms</td></tr>
<tr><td>Left ventricular ejection fraction (%)</td><td>Female: > 52%<br>Male: > 48%</td></tr>
<tr><td>Right ventricular ejection fraction (%)</td><td>Female: > 49%<br>Male: > 50%</td></tr>
<tr><td>Lungs</td><td></td><td></td></tr>
<tr><td>Lung fractional area change, right (%)</td><td>> 26%</td><td rowspan="2">Lung fractional area change, right abnormal AND/OR Lung fractional area change, left abnormal</td></tr>
<tr><td>Lung fractional area change, left (%)</td><td>> 26%</td></tr>
<tr><td>Kidney</td><td></td><td></td></tr>
<tr><td>Kidney cortical T1, right (ms)</td><td>< 1173ms</td><td rowspan="2">Kidney cortical T1, right abnormal AND/OR Kidney cortical T1, left abnormal</td></tr>
<tr><td>Kidney cortical T1, left (ms)</td><td>< 1185ms</td></tr>
<tr><td>Spleen</td><td></td><td></td></tr>
<tr><td>Spleen T1 (ms)</td><td>< 1260ms</td><td rowspan="2">Spleen T1 AND/OR spleen length abnormal</td></tr>
<tr><td>Spleen length (cm)</td><td>Female (height <164cm): < 11.1cm<br>Female (height ≥164cm): < 12.8cm<br>Male (height <177cm): < 12cm<br>Male (height ≥177cm): < 14cm</td></tr>
</table>

Table S2: Baseline characteristics of myeloid cancer patients vs healthy controls

| | **Healthy controls (n = 35)** | **Myeloid (n = 26)** | **P-value Myeloid vs. Healthy controls** |
|---|---|---|---|
| **DEMOGRAPHICS** | | | |
| **Age [mean (SD)]** | 52 (7) | 55 (13) | 0.2 |
| **Sex (Male) [n (%)]** | 12 (34%) | 13 (50%) | 0.2 |
| **BMI (Kg/m$^2$) [mean (SD)]** | 23.4 (3.1) | 27.5 (6.4) | **0.001** |
| **MULTI-ORGAN MRI METRICS** | | | |
| **Liver cT1 (ms) [mean (SD)]** | 686 (42) | 734 (93) | **0.017** |
| ≥ 800ms [n (%)] | 0 (0%) | 4 (16%) | **0.026** |
| **Liver PDFF (%) [mean (SD)]** | 2.4 (3.0) | 4.3 (4.9) | **0.01** |
| ≥ 5% [n (%)] | 2 (5.7%) | 5 (20%) | 0.12 |
| **Pancreas srT1 (ms) [mean (SD)]** | 702 (40) | 569 (50) | **<0.001** |
| ≥ 836ms [n (%)] | 0 (0%) | 0 (0%) | - |
| **Pancreas PDFF (%) [mean (SD)]** | 3.2 (2.4) | 4.4 (4.1) | 0.5 |
| ≥ 4% [n (%)] | 6 (17%) | 10 (38%) | 0.061 |
| **Mean cardiac T1 segment (ms) [mean (SD)]** | 973 (23) | 984 (48) | 0.2 |
| ≥3 cardiac segments ≥1038ms [n (%)] | 1 (2.9%) | 4 (15%) | 0.15 |
| **Mean cardiac T2 segment (ms) [mean (SD)]** | 44.6 (2.1) | 48.4 (4.0) | **0.024** |
| ≥3 cardiac segments ≥ 58ms [n (%)] | 0 (0%) | 0 (0%) | - |
| **LV ejection fraction (%) [mean (SD)]** | 60.5 (4.1) | 58.6 (6.8) | 0.18 |
| < 48% (Male) or < 52% (Female) [n (%)] | 0 (0%) | 2 (8.0%) | 1.7 |
| **RV ejection fraction (%) [mean (SD)]** | 57.8 (3.8) | 57.3 (7.7) | 0.31 |
| < 50% (Male) or < 49% (Female) [n (%)] | 1 (2.9%) | 2 (8.0%) | 0.57 |
| **Lung fractional area change, right (%) [mean (SD)]** | 45 (10) | 42 (8) | 0.12 |
| < 26% [n (%)] | 2 (5.7%) | 0 (0%) | 0.5 |
| **Lung fractional area change, left (%) [mean (SD)]** | 47 (11) | 44 (10) | 0.11 |
| < 26% [n (%)] | 2 (5.7%) | 1 (4.0%) | >0.9 |
| **Kidney cortical T1, right (ms) [mean (SD)]** | 1,061 (55) | 1,145 (75) | **<0.001** |
| ≥ 1173ms [n (%)] | 0 (0%) | 7 (28%) | **0.001** |
| **Kidney cortical T1, left (ms) [mean (SD)]** | 1,072 (51) | 1,135 (81) | **0.002** |
| ≥ 1185ms [n (%)] | 0 (0%) | 6 (24%) | **0.004** |
| **Spleen T1 (ms) [mean (SD)]** | 1,125 (71) | 1,093 (85) | 0.3 |
| ≥ 1260ms [n (%)] | 1 (2.9%) | 0 (0%) | >0.9 |
| **Spleen length (cm) [mean (SD)]** | 9.1 (2.0) | 12.8 (3.6) | **<0.001** |
| Splenomegaly [n (%)] | 2 (5.7%) | 11 (44%) | **<0.001** |
| **ORGAN ABNORMAILITY** | | | |
| **Liver abnormal [n (%)]** | 2 (5.7%) | 5 (20%) | 0.12 |
| **Pancreas abnormal [n (%)]** | 6 (17%) | 10 (38%) | 0.061 |
| **Heart abnormal (Myocarditis) [n (%)]** | 0 (0%) | 3 (12%) | 0.07 |

| | **Healthy controls (n = 35)** | **Myeloid (n = 26)** | **P-value Myeloid vs. Healthy controls** |
|---|---|---|---|
| **Lung abnormal [n (%)]** | 2 (5.7%) | 1 (4.0%) | >0.9 |
| **Kidney abnormal [n (%)]** | 0 (0%) | 7 (28%) | **0.002** |
| **Spleen abnormal [n (%)]** | 3 (8.6%) | 11 (44%) | **0.002** |
| **≥ 1 Organ(s) abnormal [n (%)]** | 10 (29%) | 19 (73%) | **<0.001** |
| **≥ 2 Organs abnormal [n (%)]** | 2 (5.7%) | 11 (42%) | **<0.001** |

Table S3: Clinical characteristics for myeloid cancer patients

| ID | Scan delta (days) | Age (yrs) | Sex | Diagnosis | Mutations | Current Treatment for myeloid neoplasm | Hb (g/L) | WCC (x10*9/L) | Plat(x10*9/L) | Neut (x10*9/L) | Creatinine (µmol/L) | eGFR (mL/min/1.73 m²) | Bili (µmol/L) | ALT (U/L) | ALP (IU/L) | MRI abnormality | Comorbidities |
|---|---|---|---|---|---|---|---|---|---|---|---|---|---|---|---|---|---|
| 002 | - | 56-60 | M | MF | CALR | No treatment | 130 | 8.39 | 411 | 5.93 | 83 | 100 | 26 | 62 | 81 | Spleen, Kidney, Liver, Pancreas & Heart | Asthma, Hypertension & Gout |
| 019 | - | 76-80 | M | MF | JAK2 | Hydroxycarbamide | 96 | 1.88 | 576 | 1.46 | 109 | 57 | 13 | 18 | 60 | Spleen & Kidney | None |
| 029 | - | 46-50 | F | MF | N/A | No treatment | *NA* | *NA* | *NA* | *NA* | *NA* | *NA* | *NA* | *NA* | *NA* | Spleen | None |
| 007 | - | 66-70 | F | ET | JAK2, TET2 | Hydroxycarbamide | 142 | 6.46 | 426 | 4.31 | 66 | 77 | 9 | 16 | 59 | None | Hypothyroidism |
| 010 | - | 61-65 | F | ET | CALR | Peg Interferon | 107 | 3.91 | 419 | 1.17 | 69 | 74 | 5 | 41 | 59 | Heart | Stroke |
| 014 | - | 56-60 | F | ET | JAK2 | Peg Interferon | 134 | 8.56 | 823 | 6.06 | 59 | 91 | 11 | 14 | 35 | Pancreas | Raynaud's |
| 018 | - | 51-55 | F | ET | JAK2 | No treatment | 128 | 11.02 | 712 | 8.23 | 57 | 97 | 10 | 10 | 45 | Spleen & Kidney | None |
| 020 | - | 46-50 | F | ET | N/A | No treatment | 141 | 12.02 | 771 | 8.94 | 47 | 122 | 16 | 16 | 66 | None | None |
| 022 | - | 41-45 | F | ET | CALR | No treatment | 141 | 9.67 | 540 | 6.12 | 64 | 89 | 5 | 13 | 62 | Kidney & Pancreas | Migraines |
| 023 | - | 46-50 | F | ET | N/A | Hydroxycarbamide | 146 | 5.83 | 508 | 4.25 | 44 | 133 | 11 | 29 | 56 | Spleen & Kidney | Coronary artery disease |
| 025 | - | 51-55 | M | ET | JAK2 | No treatment | 166 | 9.05 | 510 | 5.52 | 124 | 53 | 10 | *NA* | *NA* | Spleen & Liver | None |
| 027 | - | 31-35 | M | ET | CALR | No treatment | 145 | 6.87 | 689 | 4.35 | 85 | 90 | 14 | 101 | 89 | None | Migraines |
| 001 | - | 71-75 | M | PV | JAK2 | No treatment | 171 | 8.01 | 198 | 6.22 | 77 | 86 | 22 | 29 | 81 | Spleen, Liver & Pancreas | 2x DVTS |
| 004 | - | 56-60 | F | PV | JAK2 p | No treatment | 147 | 6.46 | 267 | 4.07 | 73 | 71 | 10 | 26 | 63 | None | Hypertension, Diverticulits & Endometriosis |
| 017 | - | 51-55 | M | PV | JAK2 | No treatment | 131 | 7.3 | 507 | 3.81 | 75 | 94 | 13 | 25 | 138 | Spleen, Liver & Pancreas | Hypothyroid, Hypertension & Type 2 Diabetes |
| 021 | - | 71-75 | M | PV | N/A | Hydroxycarbamide | 140 | 6.34 | 435 | 4.56 | 82 | 80 | 10 | 13 | 66 | Pancreas | None |
| 024 | - | 51-55 | F | PV | JAK2 | No treatment | 168 | 7.73 | 248 | 5.25 | 50 | 112 | 11 | 56 | 112 | Spleen & Liver | Haemochromatosis, |

| | | | | | | | | | | | | | | | | | |
|---|---|---|---|---|---|---|---|---|---|---|---|---|---|---|---|---|---|
| 008 | - | 71-75 | M | MDS | JAK2 PPM1D | Azacitidine | 130 | 6.06 | 902 | 3.67 | 70 | 96 | 7 | 45 | 67 | Spleen, Kidney, Pancreas & Heart | Prolactinoma & Asthma |
| 015 | - | 56-60 | M | MDS | SF3B1 TET2 | Darbepoietin | 87 | 4.35 | 338 | 2.57 | 76 | 91 | 31 | 25 | 93 | Spleen & Pancreas | Myocardial infarction |
| 003 | - | 46-50 | M | CML | N/A | Imatinib | 152 | 6.29 | 299 | 4.22 | 102 | 68 | 9 | 40 | 53 | Pancreas | None |
| 005 | - | 51-55 | F | CML | N/A | Nilotinib | 126 | 6.14 | 203 | 3.26 | 70 | 75 | 65 | 40 | 52 | Pancreas | None |
| 006 | - | 21-25 | F | CML | N/A | Dasatinib | 130 | 4.26 | 229 | 1.51 | 56 | 115 | 8 | 12 | 31 | None | Pulmonary Embolism |
| 009 | - | 41-45 | F | CML | N/A | Imatinib | 117 | 4.2 | 283 | 1.93 | 65 | 86 | 9 | 14 | 68 | Kidney | None |
| 011 | - | 61-65 | M | CML | N/A | Nilotinib | 158 | 9.8 | 286 | 6.93 | 78 | 87 | 13 | 16 | 109 | None | Gout |
| 013 | - | 41-45 | M | CML | N/A | Dasatinib | *NA* | *NA* | *NA* | *NA* | 72 | 104 | 9 | 23 | 71 | None | None |
| 016 | - | 66-70 | M | CML | N/A | Bosutinib | 124 | 8.89 | *NA* | 6.55 | 137 | 45 | 9 | 13 | 64 | Lung | Dyslipidaemia |
| TP2 | | | | | | | | | | | | | | | | | |
| 002 | 182 | 56-60 | M | MF | CALR | No change | 126 | 6.56 | 416 | 4.63 | 87 | 95 | 28 | 71 | 73 | Spleen, Kidney & Liver | No change |
| 019 | 182 | 76-80 | M | MF | JAK2 | Ruxolitinib | 94 | 5.93 | 304 | *NA* | 126 | 48 | 6 | 25 | 47 | Spleen & Kidney | No change |
| 007 | 203 | 66-70 | F | ET | JAK2 TET2 | No change | 142 | 5.31 | 427 | 3.23 | 65 | 79 | 6 | 17 | 59 | Spleen & Kidney | No change |
| 010 | 266 | 61-65 | F | ET | CALR | No change | 112 | 4.12 | 303 | 2.3 | 69 | 74 | 6 | 31 | 54 | No change | No change |
| 014 | 182 | 56-60 | F | ET | JAK2 | No change | 115 | 4.57 | 350 | 2.92 | 56 | 97 | 7 | 17 | 52 | Kidney, Pancreas & Lung | No change |
| 018 | 189 | 51-55 | F | ET | JAK2 | No change | 134 | 9.6 | 724 | 7.2 | 63 | 86 | 9 | 11 | 45 | Spleen & Kidney | No change |
| 020 | 245 | 46-50 | F | ET | N/A | No change | 137 | 10.33 | 757 | 7.25 | 53 | 106 | 18 | 27 | 71 | None | No change |
| 022 | 189 | 41-45 | F | ET | CALR | No change | 146 | 7.97 | 538 | 4.79 | 62 | 92 | 8 | 16 | 55 | Kidney | No change |
| 025 | 182 | 51-55 | M | ET | JAK2 | No change | 158 | 10.17 | 671 | 6.91 | 84 | 84 | 10 | 38 | 71 | Spleen & Liver | No change |
| 027 | 196 | 31-35 | M | ET | CALR | Interferon | 135 | 9.61 | 868 | 7.88 | 85 | 90 | 12 | 22 | 73 | Spleen | No change |
| 001 | 182 | 71-75 | M | PV | JAK2 | Hydroxycarbamide | 163 | 5.69 | 175 | 4.22 | 72 | 93 | 21 | 25 | 74 | Spleen, Liver, Pancreas & Lung | No change |
| 004 | 182 | 56-60 | F | PV | JAK2 | No change | 155 | 6.72 | 259 | 4.47 | 63 | 84 | 18 | 28 | 67 | None | No change |
| 017 | 182 | 51-55 | M | PV | JAK2 | No change | 140 | 9.8 | 502 | 6.17 | 67 | 107 | 11 | 25 | 196 | Spleen, Liver & Pancreas | No change |

| | | | | | | | | | | | | | | | | | |
|---|---|---|---|---|---|---|---|---|---|---|---|---|---|---|---|---|---|
| 021 | 217 | 71-75 | M | PV | N/A | No change | 138 | 7.33 | 418 | 5.47 | 75 | 89 | 7 | 15 | 68 | None | No change |
| 008 | 189 | 71-75 | M | MDS | JAK2 PPM1D | No change | 121 | 7.93 | 504 | 4.74 | 75 | 89 | 11 | 18 | 59 | Spleen, Kidney & Pancreas | Atrial fibrillation and coronary artery thrombus |
| 015 | 196 | 56-60 | M | MDS | SF3B1 TET2 | No change | 89 | 4.98 | 269 | 3.18 | 73 | 96 | 29 | 28 | 118 | Spleen | No change |
| 003 | 182 | 46-50 | M | CML | N/A | No change | 150 | 6.98 | 324 | 3.85 | 103 | 67 | 6 | 47 | 45 | Pancreas | No change |
| 005 | 182 | 51-55 | F | CML | N/A | No change | 123 | 6.11 | 214 | 3.01 | 127 | 38 | 49 | 26 | 57 | None | No change |
| 011 | 301 | 61-65 | M | CML | N/A | No change | 151 | 10.76 | 307 | 7.75 | 80 | 85 | 6 | 22 | 105 | None | No change |
| 013 | 266 | 41-45 | M | CML | N/A | No change | 147 | 4.88 | 247 | 2.93 | 96 | 75 | 8 | 27 | 77 | Kidney & Pancreas | No change |
| 016 | 182 | 66-70 | M | CML | N/A | No change | 123 | 8.63 | 244 | 6.18 | 137 | 45 | 8 | 12 | 87 | None | No change |

Table S4: Characteristics of myeloid cancer patients with kidney fibroinflammation

| | **Normal kidney (n = 18)** | **Abnormal kidney (n = 7)** | **P-value: Normal vs. Abnormal kidney** |
|---|---|---|---|
| **DEMOGRAPHICS** | | | |
| **Age [mean (SD)]** | 54 (13) | 56 (14) | >0.9 |
| **Sex (Male) [n (%)]** | 9 (50%) | 3 (43%) | >0.9 |
| **BMI (Kg/m$^2$) [mean (SD)]** | 27.2 (4.0) | 27.5 (10.8) | 0.3 |
| Categories [n (%)] | | | 0.5 |
| Lean (BMI < 25 Kg/m$^2$) | 5 (28%) | 4 (57%) | |
| Overweight (BMI 25 - 30 Kg/m$^2$) | 9 (50%) | 2 (29%) | |
| Obese (BMI ≥ 30 Kg/m$^2$) | 4 (22%) | 1 (14%) | |
| **MYELOID DIAGNOSIS** | | | |
| **Disease diagnosis** | | | |
| CML | 5 (28%) | 1 (14%) | 0.6 |
| MDS | 1 (5.6%) | 1 (14%) | 0.5 |
| MF | 1 (5.6%) | 2 (29%) | 0.2 |
| ET | 6 (33%) | 3 (43%) | 0.7 |
| PV | 5 (28%) | 0 (0%) | 0.3 |
| **BLOOD BIOMARKERS** | | | |
| **Hb (g/L) [mean (SD)]** | 140 (22) | 127 (17) | 0.094 |
| Categories [n (%)] | | | 0.596 |
| <130 g/L [male], <120 g/L [female] | 2 (12%) | 2 (29%) | |
| >=130 & <170 g/L [male], >=120 & <150 g/L [female] | 12 (75%) | 5 (71%) | |
| >170 g/L [male], >150 g/L [female] | 2 (12%) | 0 (0%) | |
| **WCC (x10*9/L) [mean (SD)]** | 7.10 (2.12) | 6.72 (3.19) | 0.616 |
| Categories [n (%)] | | | 0.352 |
| <4 x10*9/L | 1 (6.2%) | 1 (14%) | |
| 4 - 11 x10*9/L | 14 (88%) | 5 (71%) | |
| >11 x10*9/L | 1 (6.2%) | 1 (14%) | |
| **Neut (x10*9/L) [mean (SD)]** | 4.55 (1.98) | 4.51 (2.42) | 0.922 |
| Categories [n (%)] | | | 0.445 |
| <2 x10*9/L | 2 (12%) | 2 (29%) | |
| 2 - 7 x10*9/L | 13 (81%) | 4 (57%) | |
| >7 x10*9/L | 1 (6.2%) | 1 (14%) | |
| **Lymph (x10*9/L) [mean (SD)]** | 1.77 (0.45) | 1.50 (0.74) | 0.504 |
| Categories [n (%)] | | | 0.304 |
| <1 x10*9/L | 0 (0%) | 1 (14%) | |
| 1 - 4 x10*9/L | 16 (100%) | 6 (86%) | |
| >4 x10*9/L | 0 (0%) | 0 (0%) | |
| **Platelets (x10*9/L) [mean (SD)]** | 416 (200) | 562 (201) | 0.103 |
| Categories [n (%)] | | | 0.176 |
| <150 x10*9/L | 0 (0%) | 0 (0%) | |
| 150 - 400 x10*9/L | 8 (50%) | 1 (14%) | |

| | Normal kidney (n = 18) | Abnormal kidney (n = 7) | P-value: Normal vs. Abnormal kidney |
|---|---|---|---|
| >400 x10*9/L | 8 (50%) | 6 (86%) | |
| **Creatinine (μmol/L) [mean (SD)]** | 74 (18) | 70 (21) | 0.465 |
| Categories [n (%)] | | | 0.194 |
| <59 μmol/L [male], <45 μmol/L [female] | 0 (0%) | 1 (14%) | |
| 59 - 104 μmol/L [male], 45 - 84 μmol/L [female] | 16 (94%) | 5 (71%) | |
| >104 μmol/L [male], >84 μmol/L [female] | 1 (5.9%) | 1 (14%) | |
| **eGFR (ml/min/{1.73 m2}) [mean (SD)]** | 88 (18) | 94 (22) | 0.383 |
| Categories [n (%)] | | | 0.507 |
| <60 (ml/min/{1.73 m²}) | 1 (5.9%) | 1 (14%) | |
| ≥60 (ml/min/{1.73 m²}) | 16 (94%) | 6 (86%) | |
| **Systolic blood pressure (SBP, mmHg) [mean (SD)]** | 132 (18) | 129 (18) | 0.856 |
| Categories [n (%)] | | | >0.999 |
| SBP < 140mmHg | 14 (78%) | 5 (71%) | |
| SBP ≥ 140mmHg | 4 (22%) | 2 (29%) | |
| **Diastolic blood pressure (SBP, mmHg) [mean (SD)]** | 75 (12) | 68 (7) | 0.161 |
| Categories [n (%)] | | | >0.999 |
| DBP < 90mmHg | 16 (94%) | 7 (100%) | |
| DBP ≥ 90mmHg | 1 (5.9%) | 0 (0%) | |
| **MULTI-ORGAN MRI METRICS** | | | |
| **Liver cT1 (ms) [mean (SD)]** | 733 (94) | 730 (104) | 0.7 |
| ≥ 800ms [n (%)] | 3 (18%) | 1 (14%) | >0.9 |
| **Liver PDFF (%) [mean (SD)]** | 4.4 (4.7) | 4.4 (6.1) | 0.8 |
| ≥ 5% [n (%)] | 4 (24%) | 1 (14%) | >0.9 |
| **Pancreas srT1 (ms) [mean (SD)]** | 551 (35) | 609 (63) | **0.028** |
| ≥ 836ms [n (%)] | 0 (0%) | 0 (0%) | - |
| **Pancreas PDFF (%) [mean (SD)]** | 4.2 (3.2) | 5.6 (6.2) | >0.9 |
| ≥ 4% [n (%)] | 7 (39%) | 3 (43%) | >0.9 |
| **Mean cardiac T1 segment (ms) [mean (SD)]** | 981 (38) | 991 (68) | 0.3 |
| ≥3 cardiac segments ≥1038ms [n (%)] | 2 (11%) | 2 (29%) | 0.55 |
| **Mean cardiac T2 segment (ms) [mean (SD)]** | 47.6 (4.1) | 50.2 (3.4) | 0.14 |
| ≥3 cardiac segments ≥ 58ms [n (%)] | 0 (0%) | 0 (0%) | - |
| **LV ejection fraction (%) [mean (SD)]** | 60.3 (6.1) | 54.4 (7.2) | 0.13 |
| < 48% (Male) or < 52% (Female) [n (%)] | 1 (5.6%) | 1 (14%) | 0.49 |
| **RV ejection fraction (%) [mean (SD)]** | 59 (7) | 53 (8) | 0.33 |
| < 50% (Male) or < 49% (Female) [n (%)] | 0 (0%) | 2 (29%) | 0.07 |
| **Lung fractional area change, right (%) [mean (SD)]** | 41 (8) | 43 (9) | 0.5 |
| < 26% [n (%)] | 0 (0%) | 0 (0%) | - |
| **Lung fractional area change, left (%) [mean (SD)]** | 42 (9) | 50 (9) | 0.065 |

| | Normal kidney (n = 18) | Abnormal kidney (n = 7) | P-value: Normal vs. Abnormal kidney |
|---|---|---|---|
| < 26% [n (%)] | 0 (0%) | 0 (0%) | - |
| **Kidney cortical T1, right (ms) [mean (SD)]** | 1,108 (49) | 1,240 (29) | **<0.001** |
| ≥ 1173ms [n (%)] | 0 (0%) | 7 (100%) | **<0.001** |
| **Kidney cortical T1, left (ms) [mean (SD)]** | 1,094 (42) | 1,240 (57) | **<0.001** |
| ≥ 1185ms [n (%)] | 0 (0%) | 6 (86%) | **<0.001** |
| **Spleen T1 (ms) [mean (SD)]** | 1,101 (81) | 1,091 (95) | 0.9 |
| ≥ 1251ms [n (%)] | 0 (0%) | 0 (0%) | - |
| **Spleen length (cm) [mean (SD)]** | 12.57 (3.71) | 13.68 (3.33) | 0.3 |
| Splenomegaly [n (%)] | 6 (35%) | 5 (71%) | 0.2 |
| **ORGAN ABNORMAILITY** | | | |
| **Liver abnormal [n (%)]** | 4 (24%) | 1 (14%) | >0.9 |
| **Pancreas abnormal [n (%)]** | 7 (39%) | 3 (43%) | >0.9 |
| **Heart abnormal (Myocarditis) [n (%)]** | 1 (5.6%) | 2 (29%) | 0.18 |
| **Lung abnormal [n (%)]** | 0 (0%) | 0 (0%) | - |
| **Spleen abnormal (Splenomegaly) [n (%)]** | 6 (35%) | 5 (71%) | 0.2 |
| **≥ 1 Organ(s) abnormal [n (%)]** | 11 (61%) | 7 (100%) | 0.13 |
| **≥ 2 Organs abnormal [n (%)]** | 5 (28%) | 6 (86%) | **0.021** |
